# AI-assisted CT imaging analysis for COVID-19 screening: Building and deploying a medical AI system in four weeks

**DOI:** 10.1101/2020.03.19.20039354

**Authors:** Shuo Jin, Bo Wang, Haibo Xu, Chuan Luo, Lai Wei, Wei Zhao, Xuexue Hou, Wenshuo Ma, Zhengqing Xu, Zhuozhao Zheng, Wenbo Sun, Lan Lan, Wei Zhang, Xiangdong Mu, Chenxin Shi, Zhongxiao Wang, Jihae Lee, Zijian Jin, Minggui Lin, Hongbo Jin, Liang Zhang, Jun Guo, Benqi Zhao, Zhizhong Ren, Shuhao Wang, Zheng You, Jiahong Dong, Xinghuan Wang, Jianming Wang, Wei Xu

## Abstract

The sudden outbreak of novel coronavirus 2019 (COVID-19) increased the diagnostic burden of radiologists. In the time of an epidemic crisis, we hoped artificial intelligence (AI) to help reduce physician workload in regions with the outbreak, and improve the diagnosis accuracy for physicians before they could acquire enough experience with the new disease. Here, we present our experience in building and deploying an AI system that automatically analyzes CT images to detect COVID-19 pneumonia features. Different from conventional medical AI, we were dealing with an epidemic crisis. Working in an interdisciplinary team of over 30 people with medical and / or AI background, geographically distributed in Beijing and Wuhan, we were able to overcome a series of challenges in this particular situation and deploy the system in four weeks. Using 1,136 training cases (723 positives for COVID-19) from five hospitals, we were able to achieve a sensitivity of 0.974 and specificity of 0.922 on the test dataset, which included a variety of pulmonary diseases. Besides, the system automatically highlighted all lesion regions for faster examination. As of today, we have deployed the system in 16 hospitals, and it is performing over 1,300 screenings per day.

---

COVID-19 started to spread in January 2020. Up to early March 2020, it has infected over 100,000 people worldwide ^1^. COVID-19 causes acute respiratory distress syndrome on patients ^2,3^. Chest computed tomography (CT) imaging was shown to be an essential exam for early diagnosis. In this research, we present our experience in developing and deploying an artificial intelligence (AI) based method to assist novel coronavirus pneumonia screening using CT imaging.

Our goal is to solve two key challenges in COVID-19 screening: 1) The sudden outbreak of COVID-19 overwhelmed health care facilities in the Wuhan area. Hospitals in Wuhan had to invest significant resources to screen suspected patients, further increasing the burden of radiologists. As Ji *et al*. ^4^ pointed out, there was a significant positive correlation between COVID-19 mortality and health-care burden. It was essential to reduce the workload of clinicians and radiologists and enable patients to get early diagnoses and timely treatments. 2) In a large country like China, it is nearly impossible to train such a large number of experienced physicians in time to screen this novel disease, especially in regions without an outbreak yet. AI systems could be a good solution to both.

Specifically, we focused on helping screen COVID-19 pneumonia using chest CT imaging. The preliminary prospective analysis by Huang et al. ^2^ showed that all 41 patients in the study had abnormal chest CT, with bilateral ground-glass shape lung opacities in subpleural areas of the lungs. Many recent studies ^5–9^ also viewed chest CT as a low-cost, accurate and efficient method for novel coronavirus pneumonia diagnosis. The official guidelines for COVID-19 diagnosis and treatment (7th edition) by China’s National Health Commission ^10^ also listed chest CT result as one of the main clinical features.

Building a practical AI for epidemic response is different from typical AI diagnostic systems for the following reasons: 1) We need to build the system in days, but training AI models requires a lot of positive samples. However, at the beginning of a new epidemic, there were not many positive cases confirmed by nucleic acid test (NAT); 2) All imaging data should come from the COVID-19 patients confirmed by NAT who underwent lung CT scans. This requirement also ensured that the image data had the diagnostic characteristics; 3) Even with enough samples, training a model can require extensive annotations from experienced radiologists, but there is no reason to burden these exhausted radiologists with extra work; 4) While it is easy to distinguish pneumonia from healthy cases, it is non-trivial for the model to distinguish COVID-19 from other pulmonary diseases, which is the top clinical requirement; 5) AI systems usually require professional deployment service on-premise of the hospitals, which is not feasible in quarantine.

As illustrated in Figure 1, we designed our method to solve the above challenges: 1) Taking advantage of the end-to-end deep neural network models, we were able to adapt the models we had developed previously for other diagnoses, such as ResNet-50 ^11^ and 3D Unet++ ^12^, and construct a training-inference pipeline, evaluating a variety of candidate models quickly. 2) For the training data, in addition to the positive cases, we carefully assembled a set of negative images of inflammatory and neoplastic pulmonary diseases, such as lobar pneumonia, lobster pneumonia, and old lesions. Thus the model could learn the different features of COVID-19 from others. We also got image samples from 5 different hospitals (Table S1) with 11 different models of CT equipments (Table S2) to increase the model’s generalization ability. 3) We developed a three-stage annotation and quality control pipeline, allowing inexperienced data annotators to work with senior radiologists to create accurate annotations, with minimal time investment from the radiologists. 4) We got reasonable training result with only 131 positive cases. Then as more data came in, we retrained the model to improve the model accuracy continuously. 5) To make the diagnosis more intuitive to the radiologists and physicians, in addition to the classification model (output positive / negative predictions), we also introduced a segmentation model that highlighted lesion regions for further examination. 6) The entire tool was delivered as a low-cost, plug-and-play device, so the hospital IT staff could set it up in a self-service way, and we could remotely upgrade the models.

**Figure 1:**
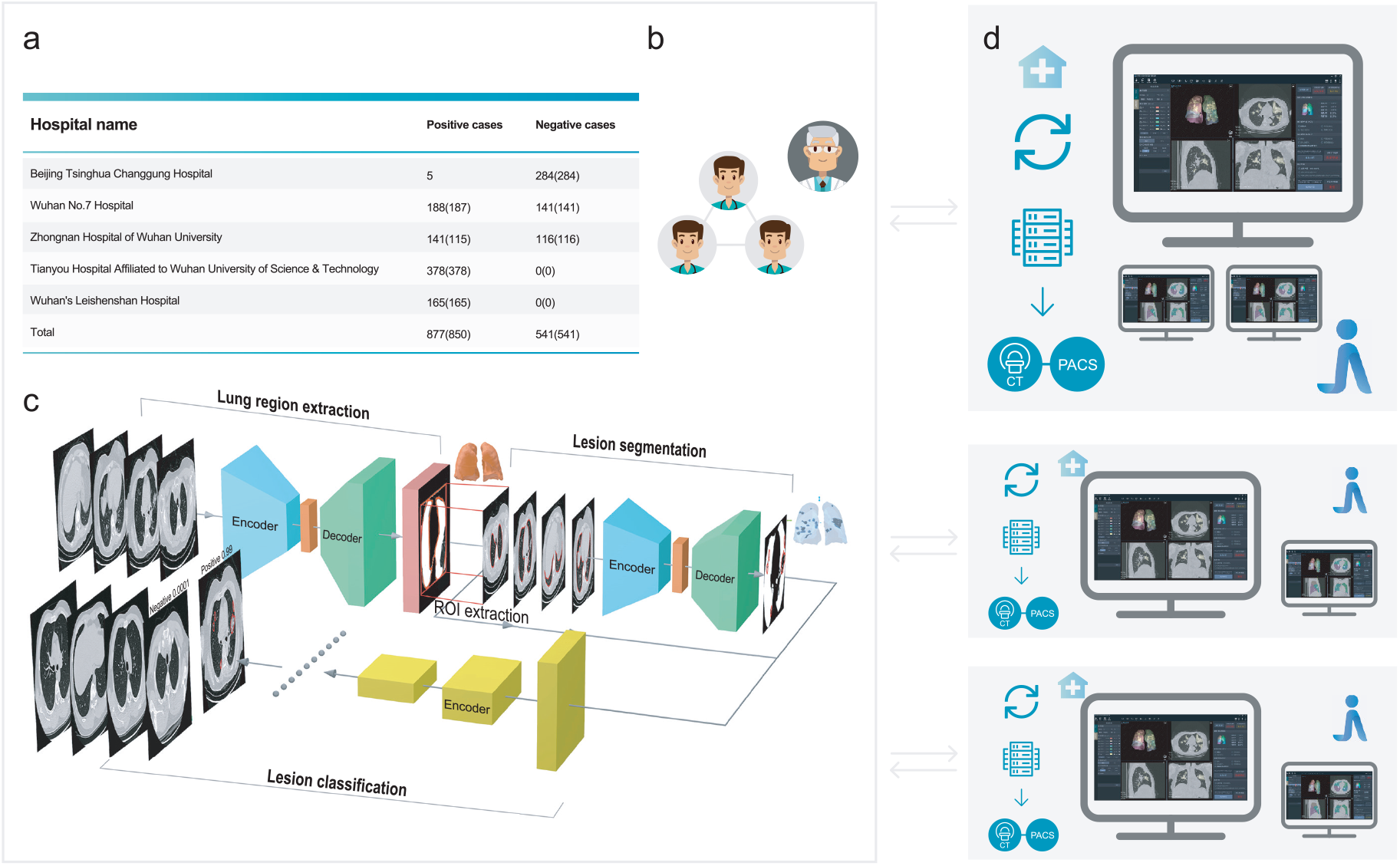
The framework of our research. **a**, Data were collected from 5 hospitals. **b**, Three-stage annotation and quality control. **c**, Deep learning model training pipeline. **d**, Deployments at hospitals.

We proposed a combined “segmentation - classification” model pipeline, which highlighted the lesion regions in addition to the screening result. The model pipeline was divided into two stages: 3D segmentation and classification. The pipeline leveraged the model library we had previously developed. This library contained the state-of-the-art segmentation models such as fully convolutional network (FCN-8s) ^13^, U-Net ^14^, V-Net ^15^, and 3D U-Net++ ^12^, as well as classification models like dual path network (DPN-92) ^16^, Inception-v3 ^17^, residual network (ResNet-50) ^11^, and Attention ResNet-50 ^18^. We selected the best diagnosis model by empirically training and evaluating the models within the library.

The latest segmentation model was trained on 732 cases (704 contained inflammation or tumors). The 3D U-Net++ model obtained the highest Dice coefficient of 0.754, and Table S3 showed the detailed segmentation model performance. By fixing the segmentation model as 3D U-Net++, we used 1,136 (723 were positive) / 282 cases (154 were positive) to train / test the classification / combined model, the detailed data distribution was given in Tables S4, S5 and S6. Figure 2(a) showed the receiver operating characteristic (ROC) curves of these four combined models. The “3D Unet++ - ResNet-50” combined model achieved the best area under the curve (AUC) of 0.991. Figure 2(a) plotted the best model by a star, which achieved a sensitivity of 0.974 and specificity of 0.922.

**Figure 2:**
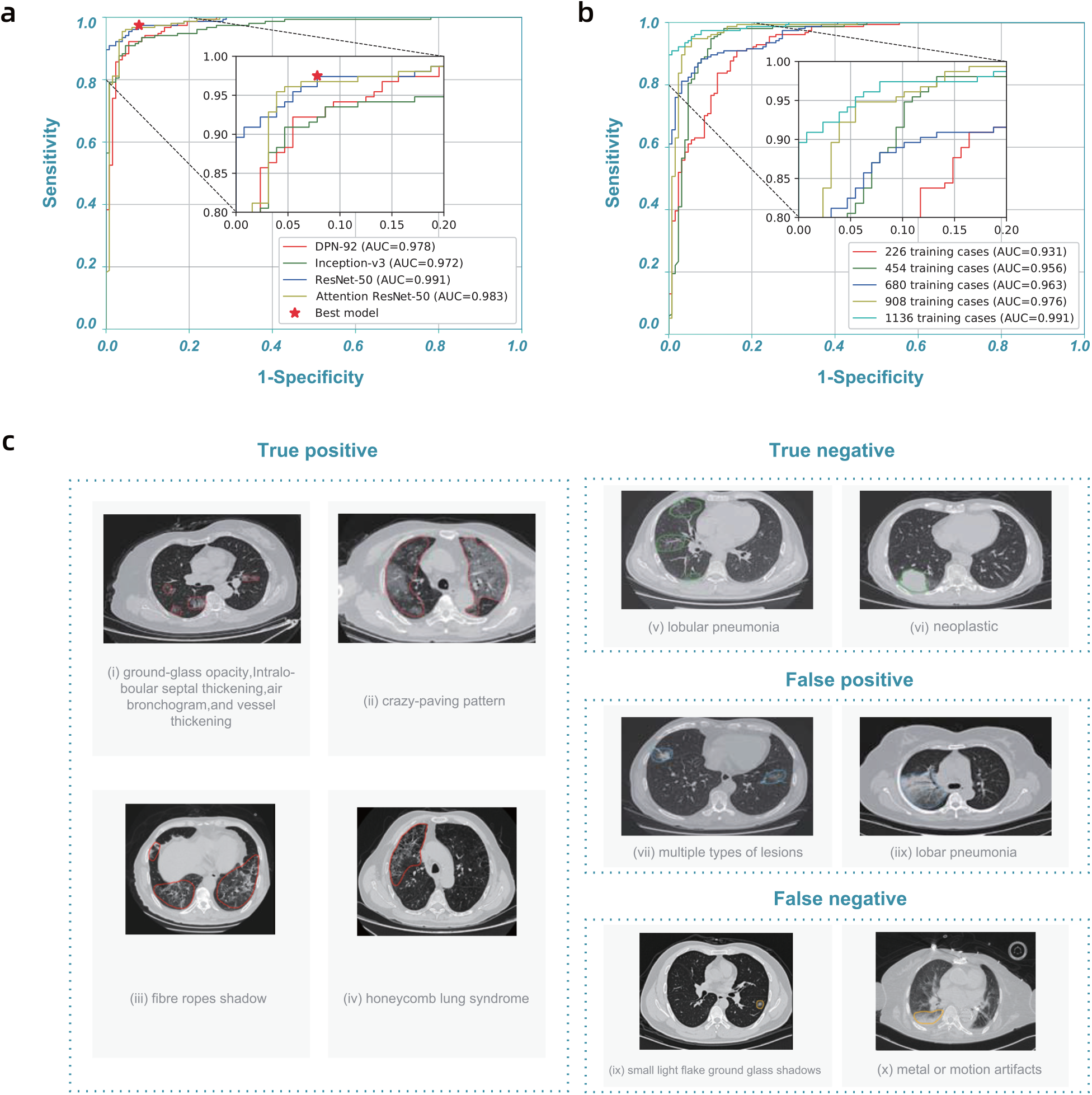
Model performance and highlights of model predictions. **a**, Receiver operating characteristic (ROC) curves of DPN-92, Inception-v3, ResNet-50, and Attention ResNet-50 with 3D U-Net++, respectively. **b**, ROC curves of 3D U-Net++ - ResNet-50 trained with different numbers of training cases. **c**, Typical predictions of the segmentation model.

The performance of the model improved steadily as the training data accumulated. In practice, the model was continually retrained in multiple stages (the average time between stages was about three days). Table S7 showed the training datasets we used in each stage. Figure 2(b) showed the improvement of the ROC curves at each stage. At the first stage, the AUC reached 0.931 using 226 training cases. The model performance at the last stage, AUC reached 0.991 with 1,136 training cases which was sufficient for clinical applications.

With the model prediction, physicians could acquire insightful information from highlighted lesion regions in the user interface. Figure 2(c) showed some examples. The model identified typical lesion characteristics of COVID-19 pneumonia, including ground-glass opacity, intralobular septal thickening, air bronchogram sign, vessel thickening, crazy-paving pattern, fibre stripes, and honeycomb lung syndrome. The model also picked out abnormal regions for cases with negative classification, such as lobular pneumonia and neoplastic lesion. These highlights helped physicians to quickly locate the slides and regions for detailed examination, improving their diagnosis efficiancy.

It was necessary to study the false positive and false negative predictions, given in Figure 2(c). Most notably, the model sometimes missed positive cases for patchy ground glass opacities with diameters less than 1 cm. The model might also introduce false positives with other types of viral pneumonia, for instance, lobar pneumonia, with similar CT features. Also, the model did not perform well when there were multiple types of lesions, or with significant metal or motion artifacts. We plan to obtain more cases with these features for training as our next steps.

At the time of writing, we had deployed the system in 16 hospitals, including Zhongnan Hospital of Wuhan University, Wuhan’s Leishenshan Hospital, Beijing Tsinghua Changgung Hospital, and Xi’an Gaoxin Hospital, etc. Physicians first ran the system once automatically, which took 0.8 seconds on average. The model prediction would be checked in the next step. Regardless of whether the classification was positive or negative, the physicians would check the segmentation results to quickly locate the suspected legions and examine if there were missing ones. Finally, physicians confirmed the screening result.

The system would help the heavily affected areas, where enough radiologists were unavailable, by giving out the preliminary CT results to speed up the filtering process of COVID-19 suspected patients. For the less affected area, it could help less-experienced radiologists, who faced a challenge in distinguishing COVID-19 from normal pneumonia, to better detect the highly-indicative features of the presence of COVID-19.

While it was not currently possible to build a general AI that could automatically diagnose every new disease, we could have a generally applicable methodology that allowed us to quickly construct a model targeting a specific one, like COVID-19. The methodology not only included a library of models and training tools, but also the process for data collection, annotation, testing, user interaction design, and clinical deployment. Based on this methodology, we were able to produce the first usable model in 7 days after we received the first batch of data, and conducted additional four iterations in the model in the next 13 days while deploying it in 16 hospitals. The model was performing more than 1,300 screenings per day at the time of writing.

Being able to take in more data continuously was an essential feature for epidemic response. The performance could be quickly improved by updating the model with continuous data taken in. To further improve the detection accuracy, we need to focus on adding training samples with complicated cases, such as cases with multiple lesion types. Besides, CT is only one of the factors for the diagnosis. We are building a multi-modal model allowing other clinical data inputs, such as patient profiles, symptoms, and lab test results, to produce a better screening result.

## Methods

### Overview

The construction of the AI model included four stages: 1) Data collection; 2) Data annotation; 3) Model training and evaluation, and 4) Model deployment. As we accumulated data, we iterated through the stages to continuously improve model performance.

### Data collection

Our dataset was obtained from 5 hospitals (see Table S1). Most of the 877 positive cases were from hospitals in Wuhan, while half of the 541 negative cases were from hospitals in Beijing. Our positive samples were all collected from confirmed patients, following China’s national diagnostic and treatment guidelines at the time of the diagnosis, which required positive results in NAT. The positive cases offered a good sample of confirmed cases in Wuhan, covering different age and gender groups (see Figure S1). We carefully chose negative cases so that in addition to healthy cases, we also had 450 cases with other known lung diseases with CT imaging features similar to COVID-19 to some extent (see Figure S2).

The hospitals used different models of CT equipment from different manufacturers (see Table S2). Due to the shortage of CT scanners for hospitals in Wuhan, slice thicknesses varied from 0.625 mm to 10 mm, with a majority (81%) under 2 mm. We believed this variety helped to improve the generalizability of our model in real deployment. In addition, we removed the personally identifiable information (PII) from all CT scans for patients’ privacy. We randomly divided the whole dataset into a training set and a test set for each model training (see Table S4).

### Data annotation

To train the models, a team of six data annotators annotated the lesion regions (if there are any), lung boundaries, and the parts of the lungs for transverse section layers in all CT samples.

Saving time for radiologists was essential during the epidemic outbreak, so our data annotators performed most of the tasks, and we relied on a three-step quality inspection process to achieve reasonable accuracy for annotation. All of the annotators had radiology background, and we conducted a four-day hands-on training led by a senior radiologist with clinical experience of COVID-19 before they performed annotations.

Our three-step quality inspection process was the key to obtaining high-quality annotations. We divided the six-annotator team into a group of four (Group A) and a group of two (Group B).

**Step 1** Group A made all the initial annotations, and Group B performed a back-to-back quality check, *i*.*e*., each of the two members in Group B checked all the annotations independently and then compared their results. The pass rate for this initial inspection was 80%. The cases that failed to pass mainly had minor errors in small lesion region missing or the inexact boundary shape.

**Step 2** Group A revised the annotations, and then Group B rechecked the annotations. This process continued until all of them passed the back-to-back quality test within the two-people group.

**Step 3** When a batch of data was annotated and passed the first two steps, senior radiologists randomly checked 30% of the revised annotations for each batch. We observed a pass rate of 100% in this step, showing reasonable annotation quality.

Of course, there might still be errors remaining, and we relied on the model training process to tolerate these random errors.

### Pre-processing

We performed the following preprocessing steps before we used them for training and testing. 1) Since different samples had different resolutions and slice thicknesses, we first normalized them to (1, 1, 2.5) mm using standard interpolation algorithms. 2) We adjusted the window width (WW) and window level (WL) for each model, generating three image sets, each with a specific window setting. For brevity, we used the [min, max] interval format in programming for WW and WL. Specifically, we set them to [-150, 350] for the lung region segmentation model, and [-1,024, 350] for both of the lesion segmentation and classification models. 3) We first ran the lung segmentation model to extract areas of the lungs from each image and used only the extraction results in the subsequent steps. 4) We normalized all the values to the range of [0, 1]. 5) We applied typical data augmentation techniques to increase the diversity of data. For example, we randomly flipped, panned, and zoomed images for more variety, which had been shown to improve the generalization ability for the trained model.

### Model library

Our model was a combination of a segmentation model and a classification model. Specifically, we used the segmentation model to obtain the lung lesion regions, and then the classification model to determine whether it was COVID-19-like for each lesion region. We selected both models empirically by training and testing all models in our previously-developed model library.

For the segmentation task, we considered several widely-used segmentation models such as fully convolutional networks (FCN-8s) ^13^, U-Net ^14^, V-Net ^15^ and 3D U-Net++ ^12^.

FCN-8s ^13^ was a “fully convolutional” network in which all the fully connected layers were replaced by convolution layers. Thus, the input of FCN-8s could have arbitrary size. FCN-8s introduced a novel skip architecture to fuse information of multi-resolution layers. Specifically, upsampled feature maps from higher layers were combined with feature maps skipped from the encoder, to improve the spatial precision of the segmentation details.

Similar to FCN-8s, U-Net ^14^ was a variant of encoder-decoder architecture and employed skip connection as well. The encoder of U-Net employed multi-stage convolutions to capture context features, and the decoder used multi-stage convolutions to fuse the features. Skip connection was applied in every decoder stage to help recover the full spatial resolution of the network output, making U-Net more precise, and thus suitable for biomedical image segmentation.

V-Net ^15^ was a 3D image segmentation approach, where volumetric convolutions were applied instead of processing the input volumes slice-wise. V-Net adopted a volumetric, fully convolutional neural network and could be trained end-to-end. Based on the Dice coefficient between the predicted segmentation and the ground truth annotation, a novel objective function was introduced to cope with the imbalance between the number of foregrounds and background voxels.

3D U-Net++ ^12^ was an effective segmentation architecture, composed of deeply-supervised encoder and decoder sub-networks. Concretely, a series of nested, dense re-designed skip pathways connected the two sub-networks, which could reduce the semantic gap between the feature maps of the encoder and the decoder. Integrate the multi-scale information, the 3D U-Net++ model could simultaneously utilize the semantic information and the texture information to make the correct predictions. Besides, deep supervision enabled more accurate segmentation, particularly for lesion regions. Both re-designed skip pathways and deep supervision distinguished U-Net++ from U-Net, and assisted U-Net++ to effectively recover the fine details of the target objects in biomedical images. Also, allowing 3D inputs could capture inter-slice features and generate dense volumetric segmentation.

For all the segmentation models, we used patch size (*i*.*e*., the input image size to the model) of (256, 256, 128). The positive data for the segmentation models were those images with arbitrary lung lesion regions, regardless of whether the lesions were COVID-19 or not. Then the model made per-pixel predictions of whether the pixel was within the lung lesion region.

In the classification task, we evaluated some state-of-the-art classification models such as ResNet-50 ^19^, Inception networks ^17,20,21^, DPN-92 ^16^, and Attention ResNet-50 ^18^.

Residual network (ResNet) ^19^ was a widely-used deep learning model that introduced a deep residual learning framework. ResNet was composed of a number of residual blocks, the shortcut connections element-wisely combined the input features with the output of the same block. These connections could assist the higher layers to access information from distant bottom layers and effectively alleviated the gradient vanishing problem, since they backpropagated the gradient to the bottom gradient without diminishing magnitude. For this reason, ResNet was able to be deeper and more accurate. Here, we used a 50-layer model ResNet-50.

Inception families ^17, 20, 21^ had evolved a lot over time, while there was an inherent property among them, which was a split-transform-merge strategy. The input of the inception model was split into a few lower-dimensional embeddings, transformed by a set of specialized filters, and merged by concatenation. This split-transform-merge behavior of inception module was expected to approach the representational power of large and dense layers but at a considerably lower computational complexity.

Dual path network (DPN-92) ^16^ was a modularized classification network that presented a new topology of connections internally. Specifically, DPN-92 shared common features and maintained the flexibility of exploring new features via dual path architectures, and realized effective feature reuse and exploration. Compared with some other advanced classification models such as ResNet-50, DPN-92 had higher parameter efficiency and was easy to optimize.

Residual attention network (Attention ResNet) ^18^ was a classification model which adopted attention mechanism. Attention ResNet could generate adaptive attention-aware features by stacking attention modules. In order to extract valuable features, the attention-aware features from different attention modules change adaptively when layers going deeper. In that way, the meaningful areas in the images could be enhanced while the invalid information could be suppressed. We used Attention ResNet-50 in the residual attention network.

All the classification models took the input of dual-channel information, *i*.*e*., the lesion regions and their corresponding segmentation masks (obtained from the previous segmentation models) were simultaneously sent into the classification models, then gave the classification results (positive or negative).

### Model training

For neural network training, we trained all models from scratch with random initial parameters. Table S4 described the training and test data distribution of both segmentation and classification tasks. We trained the models on a server with eight Nvidia TITAN RTX GPUs using the PyTorch ^22^ framework. We used Adam optimizer with an initial learning rate of 1e–4 and learning rate decay of 5e–4.

### Deployment

We deployed the trained models on workstations that we deployed on premise of the hospitals. A typical workstation contained an Intel Xeon E5-2680 CPU, an Intel I210 NIC, two TITAN X GPUs, and 64GB RAM (see Figure S3). The server imported images from the hospital’s Picture Archiving and Communication Systems (PACS), and displayed the results iteratively. The server automatically checked for model/software updates and installed them so we could update the models remotely.

### Evaluation metrics

We used the Dice coefficient to evaluate the performance of the segmentation tasks and the area under the curve (AUC) to evaluate the performance of the classification tasks. Besides, we also analyzed the selected best classification model with sensitivity and specificity.

Concretely, the Dice coefficient was the double area of overlap divided by the total number of pixels in both images, which was widely used to measure the ability of the segmentation algorithm in medical image segmentation tasks. AUC denoted “area under the ROC curve”, in which ROC stood for “receiver operating characteristic”. ROC curve was drawn by plotting the true positive rate versus the false positive rate under different classification thresholds. Then AUC calculated the two-dimensional area under the entire ROC curve from (0,0) to (1,1), which could provide an aggregate measure of the classifier performance across varied discrimination thresholds. Sensitivity / specificity was also known as the true positive / negative rate, measured the fraction of positives / negatives that were correctly identified as positive / negative.

## Data Availability

Please contact the authors for data availability.

## Ethics

This study was approved by the Ethics Committee of Beijing Tsinghua Changgung Hospital. Written informed consent was provided by all prospective participants. For patients whose CT scans were stored in the retrospective databases, informed consent was waived by the Ethics Committee.

## Acknowledgements

This work is supported by National Key Research and Development Program of China No. 2020YFC0845500, National Natural Science Foundation of China (NSFC) No. 61532001, Tsinghua Initiative Research Program Grant No. 20151080475, Application for Independent Research Project of Tsinghua University (Project Against SARI).

## Author Contributions Statement

Z.Y., J.D., B.W. and S.J. proposed the research, X.W., J.D. and J.W. led the multicenter study, H.X., Z.Z., X.W., J.W., H.X., L.L., W.S., S.J. and L.W. collected data, X.H., B.Z., Z.Z., Z.J., C.S., M.L. and J.G. performed the data annotation, W.Z., B.W. and W.Z. wrote the deep learning code and performed the experiment, Z.Z., X.M. and W.S. evaluated the algorithm, H.J., Z.X., B.W., W.Z. and S.J. designed and developed the deployment system, W.Z., S.J., B.W., Z.X. and Z.R. deployed the equipment, W.M., C.S., Z.W., J.L., S.W., W.X., H.X., B.W. and W.S. wrote the manuscript, X.W., J.D., W.X., J.W. and L.Z. reviewed the manuscript.

